# A Mixed Methods Study of Program-Level Factors Influencing Patient and Family Engagement in First Episode Psychosis Coordinated Specialty Care

**DOI:** 10.64898/2026.01.27.26344928

**Authors:** Cheryl Y. S. Foo, Catherine J. Leonard, Merranda M. McLaughlin, Kelsey A. Johnson, Dost Öngür, Kim T. Mueser, Corinne Cather

## Abstract

**Background:** Poor patient retention and family engagement compromise the effectiveness of coordinated specialty care (CSC) for first-episode psychosis (FEP). This mixed methods study aimed to identify program-level characteristics (CSC fidelity and engagement strategies) associated with patient retention and family engagement in Massachusetts CSC programs.

**Methods:** Primary outcomes were rates of patient retention and family engagement (≥1 evidence-based family intervention session), based on CSC program census (October 2022 - September 2023). Quantitative analyses explored program characteristics (EPINET Program-Level Core Assessment Battery) and fidelity ratings (Massachusetts Psychosis Fidelity Scale) as predictors using t-tests or univariate linear regressions. Thematic analysis of program interviews compared patient and family engagement strategies employed by high versus low performing programs.

**Results:** Across nine programs, mean patient retention was 86% (range: 58-97%) and family engagement was 40% (range: 12-100%). Higher fidelity to evidence-based services (e.g., individual therapy, family intervention, and supported education/employment) was significantly associated with both outcomes (p<.05; R**^2^** range: .51-.72). Mixed-methods analysis showed that high performing programs used case management-related supports to meet service users’ practical needs. Factors associated with higher patient retention included having comprehensive intake assessments, provider visits during hospitalization, and periodic treatment reviews. Programs that conducted benefits counseling and proactively recommended family services as standard care had higher family engagement.

**Conclusions:** Higher fidelity CSC programs had better patient retention and family engagement. Case management-related supports addressed treatment barriers. Strategies designed to strengthen therapeutic alliance and goal alignment may promote patient engagement, while family engagement may benefit from proactive recommendation of family intervention.

## 1. Introduction

Early intervention after a first-episode of psychosis (FEP) can lead to long-term improvements in symptoms and psychosocial functioning (Correll et al., 2018). Coordinated specialty care (CSC) is the gold-standard, evidence-based treatment for FEP in the United States (U.S.) (Kane et al., 2016), providing recovery-oriented, team-based care including pharmacotherapy, cognitive-behavioral psychotherapy, family education and support, supported employment and education, case management (Heinssen & Azrin, 2022), and peer support (Kazandjian et al., 2022). However, poor treatment engagement is a significant challenge, with 15-50% of patients disengaging before completing the recommended two years of treatment. Most patients who end treatment prematurely disengage within the first 6-9 months (Kane et al., 2016). Treatment disengagement thus substantially compromises CSC effectiveness, putting patients at greater risk for relapse and increased long-term morbidity (Howes et al., 2021).

The underutilization of evidence-based family intervention for psychosis, a core CSC element, also undermines the promise of CSC. While most individuals enrolled in CSC express the desire for some degree of family involvement in their care and 74-84% of families have contact with the treatment team (Jones et al., 2021), only 22-46% participate in evidence-based family intervention in the U.S. (Breitborde et al., 2015; Glynn et al., 2018). This gap persists despite robust evidence that family intervention enhances patients’ recovery outcomes and decreases relapse rates by approximately 50% over treatment as usual (Claxton et al., 2017; Gleeson et al., 2025).

Research has identified individual-level predictors of FEP treatment disengagement, including longer duration of untreated psychosis (DUP), persistent substance use, poor insight, and medication non-adherence (Robson & Greenwood, 2022). However, these characteristics are either immutable (e.g., DUP) or difficult to modify early in treatment. Moreover, qualitative research has identified family-related factors (e.g., access, support needs/preferences) and organizational barriers (e.g., inadequately trained staff) as contributing to low uptake of family intervention (Eassom et al., 2014; Selick et al., 2017).

One promising strategy for improving treatment retention and family intervention uptake is to identify programmatic factors within CSC teams that can be implemented or modified. Recent research indicates that patients in programs with higher fidelity to the CSC model experienced greater symptom and social functioning improvement than those in lower-fidelity programs (Rosenblatt et al., 2025). Better engagement of patients and family members in treatment could be one of the mechanisms by which higher-fidelity CSC programs have a greater impact on outcomes. However, the contribution of program characteristics and fidelity on treatment engagement has not been studied. Additionally, most studies’ focus on barriers to patient/family involvement, however, there is limited empirical data on effective strategies for engaging patients and families. Thus, this mixed methods study aimed to identify modifiable program-level characteristics and practices associated with patient retention and family engagement rates across Massachusetts’ CSC programs.

## 2. Methods

### 2.1. Study Overview and Participating Programs

Quantitative and qualitative data were collected from nine of ten CSC programs in Massachusetts providing at least two years of services to people within three to five years of their psychotic disorder illness onset (ages: 13 to 40 years). One site declined to participate. All programs were part of the Massachusetts Psychosis Network for Early Treatment (MAPNET; https://www.mapnet.online), a statewide training and technical assistance center funded by the Massachusetts Department of Mental Health (DMH).

The primary outcomes of patient retention and family engagement rates were determined from annual program census reporting by each CSC team. Program characteristics and fidelity assessments were examined as predictors. Based on group interviews with CSC providers, strategies for engaging patients and families were identified and compared between programs with higher vs. lower rates of patient retention/family engagement.

### 2.2. Ethics

The Massachusetts General Brigham Institutional Review Board (MGB IRB) exempted the survey and interview procedures from review as data were collected for quality improvement purposes (2023P003073). The annual program census reporting and site visits were part of routine fidelity and contract monitoring through the Massachusetts DMH. Secondary data analysis of census reports and fidelity ratings was approved by the Massachusetts DMH and MGB IRBs (2019P000433).

### 2.3. Measures and Procedures

#### 2.3.1. Patient Retention and Family Engagement Rates

Based on collated monthly reports, CSC team leaders reported program census for the period of October 1, 2022, to September 30, 2023, including numbers of: 1) total active patients, 2) patients who discontinued services over the program duration (excluding successful program completion, meeting recovery goals, ineligibility (e.g., moving out of area), or death), and 3) patients whose families participated in at least one session of an evidence-based family intervention for psychosis (e.g., psychoeducation, single family therapy, multifamily group). The number of retained patients was calculated by subtracting the number of discontinued patients from the number of total active patients. Patient retention and family engagement rates were calculated by dividing the retained patients and engaged families by total census, respectively.

#### 2.3.2. Program Characteristics

Team leaders reported program characteristics using the standardized Early Psychosis Intervention Network (EPINET) Program-Level Core Assessment Battery (EPINET, 2022), including enrollment criteria, treatment model, patient travel distance, staff-to-patient ratio, funding model, CSC components provided, and the modes of CSC services provided (e.g., telehealth, in-person). Additional questions on family services covered: team leader-rated prioritization of family services (one item; 5-point Likert scale), staffing and training of family clinicians, types and formats of family interventions, family contact, and family educational materials and resources provided. Team leaders received $30 remuneration.

#### 2.3.3. Fidelity Assessment

Fidelity assessments were collected throughout 2023 by trained independent assessors from MAPNET, using The Massachusetts Psychosis Fidelity Scale (MAPS; measure available upon request) (Kline & Johnson, 2021), adapted from the widely-used and validated First Episode Psychosis Services Fidelity Scale (FEPS-FS 1.0) (Addington, 2021; Addington et al., 2024) to reflect the priority practices and intended delivery of CSC programs in Massachusetts. FEPS-FS and MAPS ratings are highly correlated (DeTore et al., under review).

We used fidelity ratings from 20 specific domains across three broad categories in MAPS: 1) Staffing and Services (seven domains); 2) Program Structure (six domains); and 3) Program Procedures (seven domains). Each domain has a set of related criteria in a checklist format. See Appendix A for full list of MAPS’ categories, domains, and number of criteria within each domain assessed in the study. For example, under the Staffing and Services category, fidelity to team leader role, individual therapy, family services, medication management, health management, supported education and employment, and case management were assessed. Under the “family services” domain, criteria included: 1) providing evidence-based family intervention; 2) receiving training in evidence-based family intervention; 3) providing ≥8 sessions of evidence-based family intervention to ≥70% families in first year of treatment; 4) having ≥60% of families involved in intake assessment, 5) having a signed release of information for family contact, and 6) having a team member contact families at least quarterly. While the FEPS-FS assesses fidelity for the family therapy domain based on the family engagement benchmark indicated in criteria 3, the MAPS expands this domain to assess the extent to which families are involved in treatment in other ways (e.g., contact with team members, involvement in assessment and treatment planning). Assessments were based on a site visit, observation of a clinical team meeting, individual meetings with providers of different CSC services, and reviews of randomly selected charts.

While the MAPS fidelity ratings use an ordinal scoring system for domain scores (i.e., 0-2), in this study, a domain score was calculated by summing the number of criteria met to yield a continuous score with a larger range to maximize variability and thereby enhance sensitivity to predicting retention/engagement rates. Engagement benchmark criteria under the “individual therapy” and “family services” domains were removed to avoid confounding with the primary outcomes of patient retention and family engagement. Overall staffing, structure, and procedure category scores were calculated by summing their domain scores, and an overall fidelity score was calculated by summing all three category scores.

#### 2.3.4. Barriers and Engagement Strategies

Semi-structured program interviews (conducted June-July 2024) identified barriers to and strategies for patient and family engagement experienced by each program. Sixty-minute, online interviews with various CSC providers (facilitated by C.Y.S.F) were audio-recorded and transcribed. Interviewees received $50 remuneration. See Appendix B for interview guide.

### 2.4. Quantitative Analysis

Program characteristics and fidelity scores (overall fidelity, category, and domain scores) were explored as predictors of patient retention and family engagement rates, respectively, using independent t-tests or univariate linear regressions. If a fidelity domain was significantly associated (p≤.05) with the outcome, independent t-tests were used to further explore effects of each criterion on the outcome. Hedges’ g, an effect size measure for small sample sizes, was calculated for each association. Effect sizes of 0.20 to 0.49, 0.50 to 0.79, and ≥ 0.80 are considered small, medium, and large, respectively (Cohen, 2009). Statistical analyses were conducted on SPSS v29.

### 2.5. Qualitative and Mixed Methods Analysis

Thematic analysis identified perceived treatment barriers and engagement strategies for patients and families, respectively. Research team members (C.Y.S.F., C.J.L., M.M.) independently coded transcripts, applying a deductive framework informed by barriers and facilitators identified in the literature (Eassom et al., 2014), as well as inductively identifying other engagement strategies. Intercoder reliability was assessed on four randomly-selected transcripts (O’Connor & Joffe, 2020) satisfactory reliability (κ≥0.75) was achieved for intercoder reliability (Campbell et al., 2013). Coding differences and codebook refinements were resolved after each round of coding to ensure that identified strategies were comprehensively and accurately captured.

To identify strategies associated with better engagement outcomes, we categorized programs by mean split into high and low patient retention and (separately) family engagement programs. Matrix coding query compared endorsement frequency of identified strategies across programs with high vs. low patient retention, and high vs. low family engagement, respectively. Qualitative and mixed methods analyses were conducted with NVivo 15.

## 3. Results

Across the nine participating programs, seven implemented the NAVIGATE program (Mueser et al., 2015), while others used PREP, a milieu-based model (Caplan et al., 2013), or OnTrack (https://www.ontrackny.org). All programs conducted hybrid telehealth/in-person visits, and four provided home/in-community visits. Eight programs provided evidence-based family intervention (e.g., NAVIGATE Family Education and Support program [n=7]; the McFarlane Multifamily Group (McFarlane, 2022) [n=3]), while one program did not provide any family services. Four programs provided a family support/process group, and three programs had family peer support services. See Appendix Table C.1. for each program’s key characteristics.

From October 2022 to September 2023, the nine CSC programs served a total of 581 patients (M: 65; SD: 33; range: 19-201). Mean patient retention was 86% (SD: 13; high patient retention teams (n=6): 89% - 97%; low retention teams (n=3): 58% - 76%). Mean family engagement was 40% (SD: 27; high family engagement teams (n=4): 49% to 100%; low engagement teams (n=5): 12% - 27%) (**Figure 1**). Patient retention and family engagement rates were non-significantly correlated (r=0.2, p=.79). Fidelity ratings across programs were high overall (M: 79.7, SD: 4.6, based on continuous MAPS scoring and out of a total score of 89). Detailed results on fidelity ratings are reported elsewhere (DeTore et al., under review).

**Figure 1.**
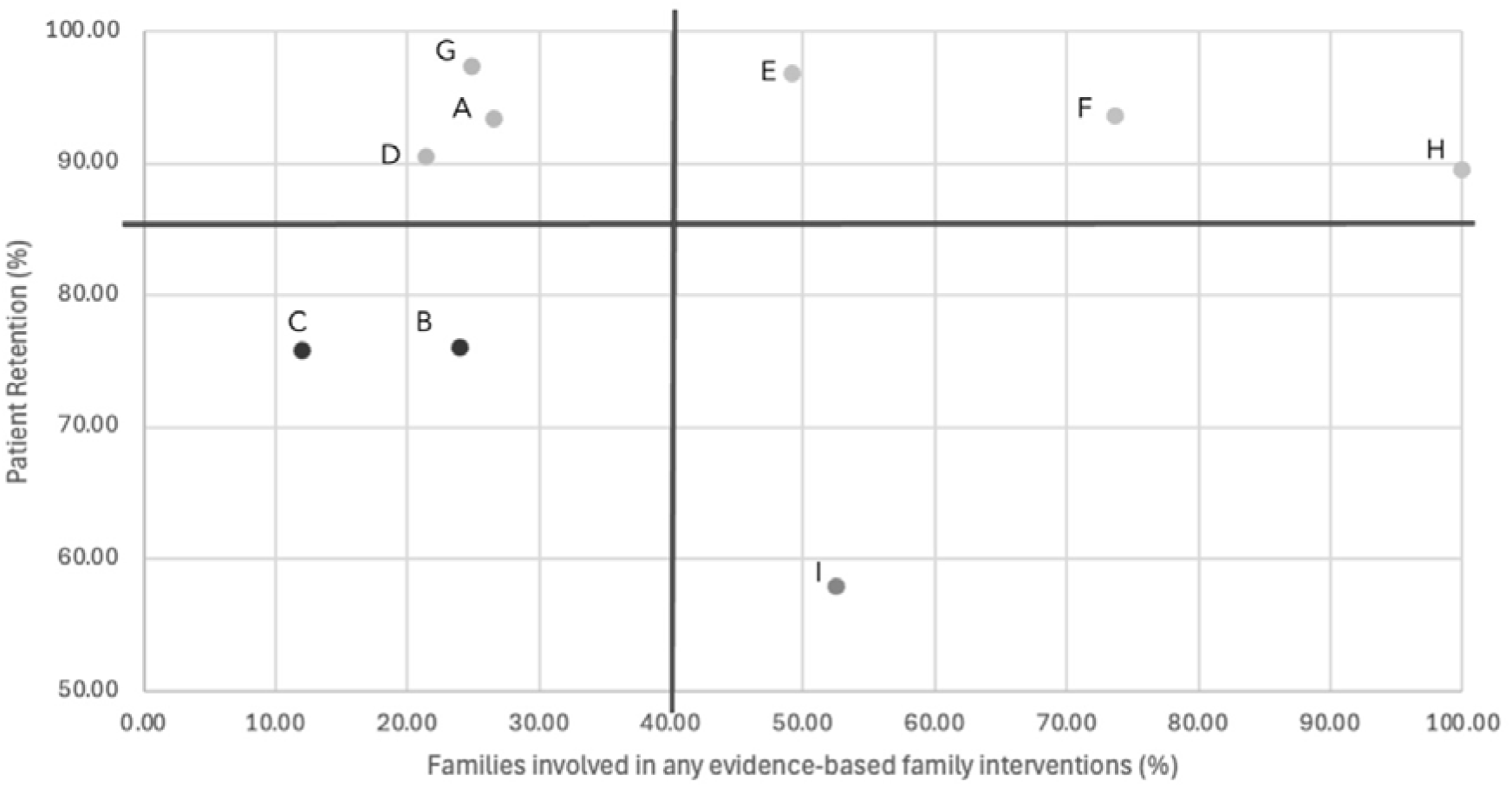
Patient Retention and Family Engagement Rates by Program. ***Caption for Figure 1:*** Figure indicates each program’s patient retention rates (y-axis) and family engagement rates (x-axis). Programs above the horizontal line (mean patient retention rate: 86%) were categorized as “higher patient retention” programs (n=6) and those below the line were “lower patient retention” programs (n=3). Programs on the left of the vertical line (mean family engagement rate: 40%) were categorized as “higher family engagement” programs (n=5) and those on the right were categorized as “lower family engagement” programs (n=4).

### 3.1. Patient Retention

#### 3.1.1. Fidelity and Program-Level Correlates

Quantitative analysis identified several program-level factors that were significantly positively associated with patient retention rates (Table 1; full list of explored predictors in Appendix Table D.1.). In terms of structural characteristics, programs (n=8) that enrolled a majority of patients with a psychotic disorder within five years of illness onset, or provided individual therapy, or family intervention, or peer support services had higher retention rates than the program that did not (p≤.01).

**Table 1.** Key Program-Level Factors Affecting Patient Retention Rates.

| Program-Level Factor | N (%) / M (SD) | Observed mean patient retention rate at min x (SD) | B (SE) | Effect size g [95% CI] / R<sup>2</sup> | p-value |
| --- | --- | --- | --- | --- | --- |
| **Services Provided** |  |  |  |  |  |
| In-clinic/telehealth family intervention | 8 (89%) | 57.9 | 31.2 (9.1) | 3.23 [0.69, 5.65] | **.01\*** |
| Peer support services | 8 (89%) | 57.9 | 31.2 (9.1) | 3.23 [0.69, 5.65] | **.01\*** |
| **Massachusetts Psychosis Fidelity Scale Assessment (Score Range)** |  |  |  |  |  |
| **Total Fidelity Score (0-89)** | 79.7 (4.6) | 97.3 | 0.8 (1.0) | .08 | .44 |
| **Total Staffing and Services Score (0-34)** | 29.8 (2.2) | 84.9 (10.7) | 1.1 (2.3) | .03 | .65 |
| **Individual Therapy (0-2)** | 2.8 (0.4) | 57.9 | 31.2 (9.9) | .62 | **.01\*** |
| - Trained in evidence-based individual therapy | 8 (89%) | 57.9 | 31.2 (9.9) | 3.23 [0.69, 5.65] | **.01\*** |
| **Family Services (0-5)** | 4.2 (1.0) | 69.9 (10.4) | 11.5 (2.7) | .72 | **<.01\*** |
| - Provides evidence-based family interventions | 8 (89%) | 57.9 | 31.2 (9.1) | 3.23 [0.69, 5.65] | **.01\*** |
| - Trained in evidence-based family interventions | 8 (89%) | 57.9 | 31.2 (9.1) | 3.23 [0.69, 5.65] | **.01\*** |
| **Total Structure Score (0-19)** | 17.7 (1.0) | 57.9 | 7.5 (4.1) | .33 | **.10** |
| 80% of patients meeting eligibility criteria | 8 (89%) | 57.9 | 31.2 (9.9) | 3.23 [0.69, 5.65] | **.01\*** |
| **Coordination with Inpatient Services (0-7)** | 6.6 (0.5) | 80.7 (8.4) | 13.3 (4.0) | .65 | **.02\*** |
| - Visit with patient on inpatient unit | 5 (56%) | 80.7 (8.4) | 13.3 (4.0) | 2.11 [0.37, 3.78] | **.02\*** |
| **Total Procedure Score (0-36)** | 32.2 (2.0) | 97.2 | 1.3 (2.4) | .04 | .61 |
| **Clinical and psychosocial needs assessment (0-11)** | 10.4 (0.7) | 57.9 | 15.9 (3.3) | .77 | **<.01\*** |
| - Individual and family psychiatric history at intake | 8 (89%) | 57.9 | 31.2 (9.1) | 3.23 [0.69, 5.65] | **.01\*** |
| - Feedback is offered to client (and family) | 5 (56%) | 75.8 (14.6) | 17.6 (6.6) | 1.58 [0.12, 2.97] | **.03\*** |

Note. $p \leq .10$ is **bolded**. \*: $p \leq .05$ . NA: Not applicable as low cell count.

Programs that delivered evidence-based individual therapy (B=31.2 (9.9), R^2^=.62, p<.01) or family services (B=11.5 (2.7), R^2^=.72, p<.01) with higher fidelity demonstrated significantly higher patient retention rates. Better care coordination with inpatient services (B=13.3 (4.0), R^2^=.65, p=.02), particularly provider visits during hospitalization (n=5, g [95% CI]=2.11 [0.37, 3.78], p=.02), was associated with higher patient retention rates. Finally, teams with more comprehensive intake assessments (B=15.9 (3.3), R^2^=.77, p<.01), and in particular those that provided specific feedback on diagnosis and recommended treatment plan to patients and family members (n=5, g=1.58 [0.12, 2.97], p=.03), were significantly associated with higher retention rates.

#### 3.1.2. Barriers and Engagement Strategies

Qualitative data indicated providers perceived several barriers to patient retention: 1) adverse social determinants of health (e.g., housing instability, transportation difficulties); 2) patient illness-related factors (e.g., poor insight, substance use); and 3) inadequate staffing to deliver all CSC components (Appendix Table E.1.). There were no differences between providers in high (n=6) versus low (n=3) patient retention programs in the identification of any of the barriers.

Providers in high and low patient retention programs both endorsed a number of engagement strategies, including: 1) building trust and rapport; 2) practicing person-centered care; 3) adopting a whole-team approach to engagement; 4) using a program orientation to set treatment expectations (prioritizing medication management and individual therapy); 5) using assertive outreach and having flexible attendance policies; and 6) using multiple treatment delivery formats (Appendix Table E.2.).

However, in contrast to the lack of difference in perceived barriers between high and low patient retention teams, there were several engagement strategies that differed between the programs (summarized in **Table 2** with relevant quotes). All high retention programs stated that they prioritized and provided case management and practical support to address treatment barriers and advocated for psychosocial supports in the community (e.g., Individualized Education Program (IEP), school placements), compared to only 33% of low retention programs. Additionally, 67% of the high retention programs reported implementing periodic treatment reviews with patients and families, a practice not reported in lower retention programs. Low retention programs instead reported using team meetings or clinical supervision to discuss treatment planning. Providers in high retention programs expressed how treatment reviews (conducted annually or in the first few months after enrollment) helped align treatment goals with CSC services to enhance treatment engagement.

**Table 2.**
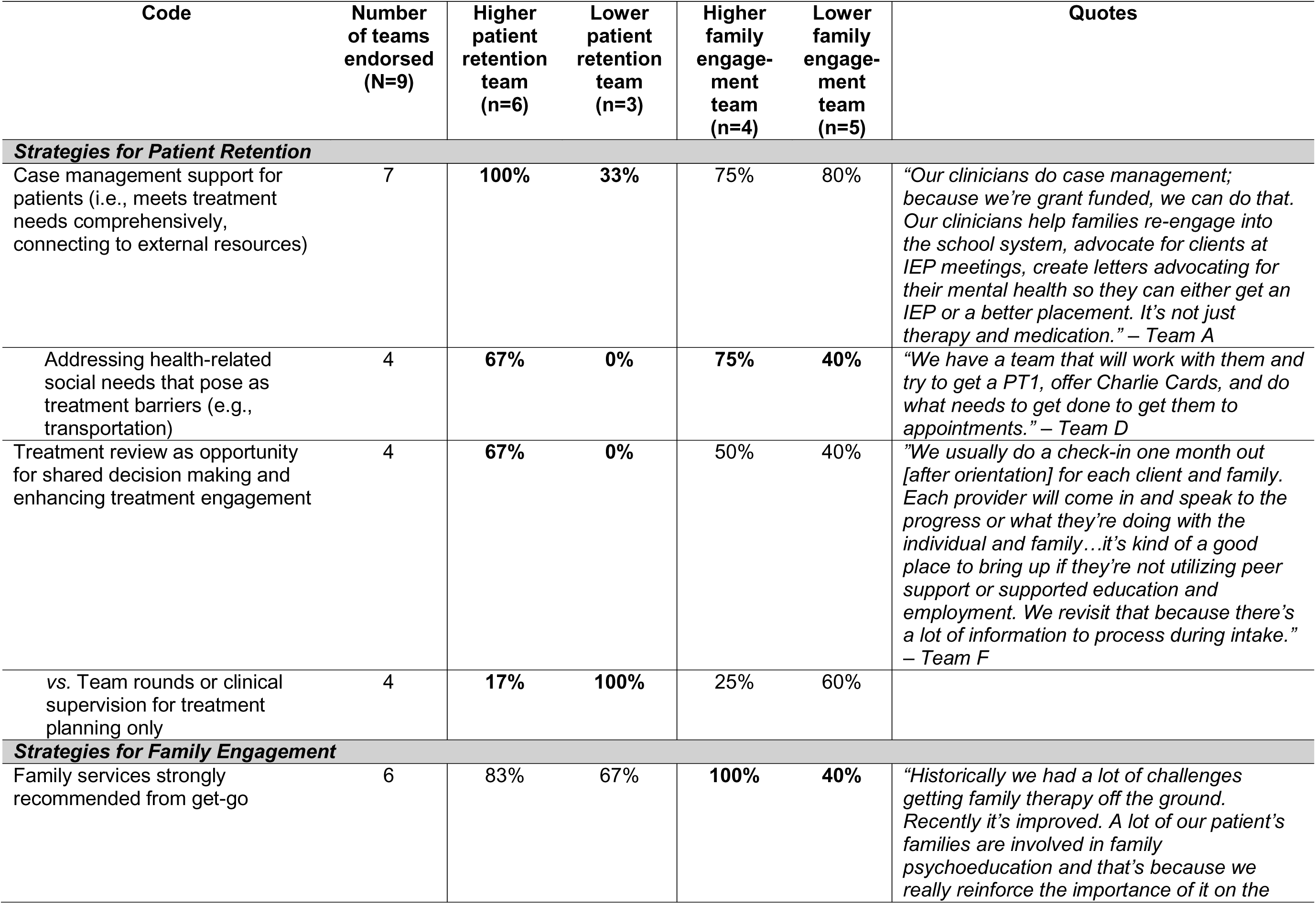
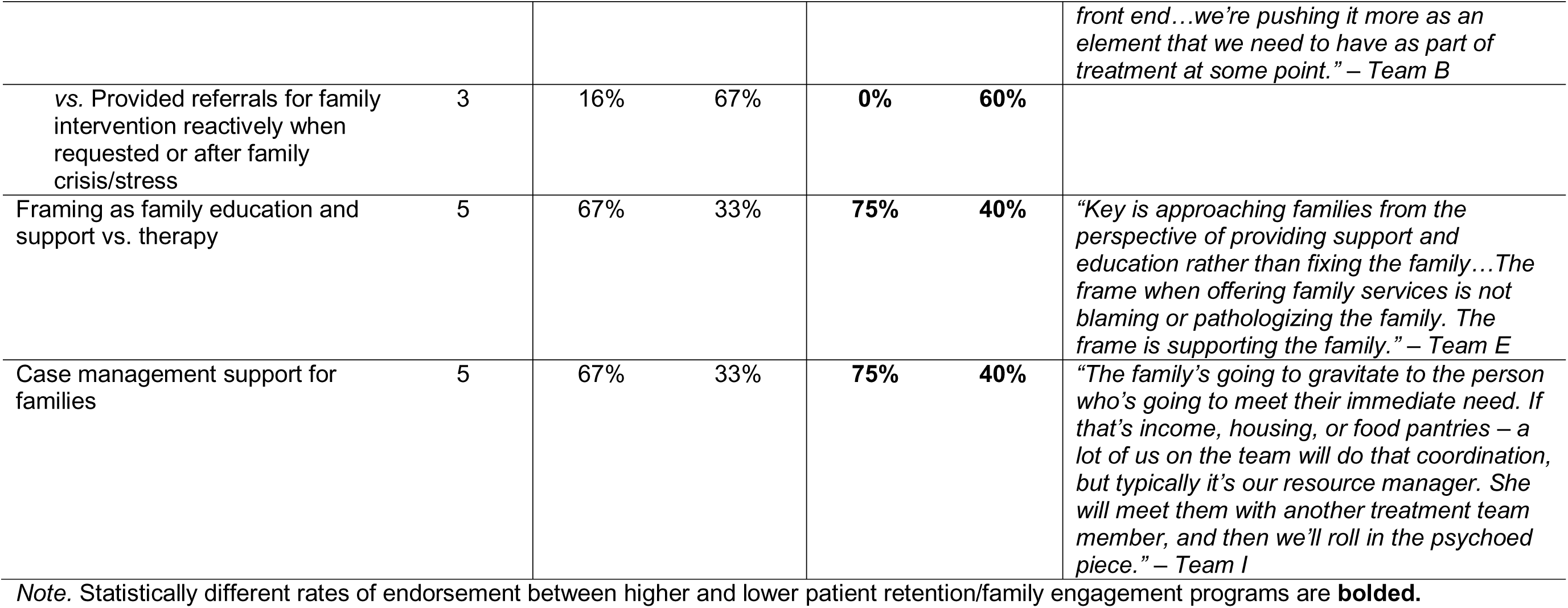
Engagement Strategies Differentiating Higher and Lower Patient Retention and Family Engagement Programs.

| Code | Number of teams endorsed (N=9) | Higher patient retention team (n=6) | Lower patient retention team (n=3) | Higher family engagement team (n=4) | Lower family engagement team (n=5) | Quotes |
| --- | --- | --- | --- | --- | --- | --- |
| **Strategies for Patient Retention** |  |  |  |  |  |  |
| Case management support for patients (i.e., meets treatment needs comprehensively, connecting to external resources) | 7 | 100% | 33% | 75% | 80% | "Our clinicians do case management; because we're grant funded, we can do that. Our clinicians help families re-engage into the school system, advocate for clients at IEP meetings, create letters advocating for their mental health so they can either get an IEP or a better placement. It's not just therapy and medication." – Team A |
| Addressing health-related social needs that pose as treatment barriers (e.g., transportation) | 4 | 67% | 0% | 75% | 40% | "We have a team that will work with them and try to get a PT1, offer Charlie Cards, and do what needs to get done to get them to appointments." – Team D |
| Treatment review as opportunity for shared decision making and enhancing treatment engagement | 4 | 67% | 0% | 50% | 40% | "We usually do a check-in one month out [after orientation] for each client and family. Each provider will come in and speak to the progress or what they're doing with the individual and family...it's kind of a good place to bring up if they're not utilizing peer support or supported education and employment. We revisit that because there's a lot of information to process during intake." – Team F |
| vs. Team rounds or clinical supervision for treatment planning only | 4 | 17% | 100% | 25% | 60% |  |
| **Strategies for Family Engagement** |  |  |  |  |  |  |
| Family services strongly recommended from get-go | 6 | 83% | 67% | 100% | 40% | "Historically we had a lot of challenges getting family therapy off the ground. Recently it's improved. A lot of our patient's families are involved in family psychoeducation and that's because we really reinforce the importance of it on the *front end...we're pushing it more as an element that we need to have as part of treatment at some point." – Team B* |
| vs. Provided referrals for family intervention reactively when requested or after family crisis/stress | 3 | 16% | 67% | **0%** | **60%** |  |
| Framing as family education and support vs. therapy | 5 | 67% | 33% | **75%** | **40%** | *"Key is approaching families from the perspective of providing support and education rather than fixing the family...The frame when offering family services is not blaming or pathologizing the family. The frame is supporting the family." – Team E* |
| Case management support for families | 5 | 67% | 33% | **75%** | **40%** | *"The family's going to gravitate to the person who's going to meet their immediate need. If that's income, housing, or food pantries – a lot of us on the team will do that coordination, but typically it's our resource manager. She will meet them with another treatment team member, and then we'll roll in the psychoed piece." – Team I* |

*Note.* Statistically different rates of endorsement between higher and lower patient retention/family engagement programs are **bolded**.

### 3.2. Family Engagement

#### 3.2.1. Fidelity and Program-Level Correlates

Several program-level factors were significantly associated with higher family engagement rates (Table 3; full list of explored predictors in Appendix Table D.2.). Programs whose team leaders strongly agreed that family services were of high priority in their program (n=5) had significantly higher family engagement rates (g=1.45 [0.03, 2.79], p=.05). Overall fidelity across CSC components (“Staffing and Services” scale) was also associated with higher family engagement (B=9.8 (2.8), R^2^=.63, p=.01). Higher fidelity supported education and employment services (B=7.9 (3.2), R^2^=.51, p=.05), and in particular programs that provided benefits counselling about social security disability benefits, Medicaid, and other government entitlements (g=1.60 [0.03, 3.55], p=.05), was associated with higher family engagement.

**Table 3.** Key Program-Level Factors Affecting Family Engagement Rates.

| Factor | N (%) / M (SD) | Observed mean family engagement rate at min x (SD) | B (SE) | Effect size g [95% CI] / AR<sup>2</sup> | p-value |
| --- | --- | --- | --- | --- | --- |
| High leadership prioritization of family services | 5 (56%) | 21.1 (6.3) | 34.3 (14.1) | 1.45 [0.03, 2.79] | **.05 \*** |
| **Services Provided** |  |  |  |  |  |
| In-community family intervention visits | 4 (44%) | 27.1 (15.2) | 29.4 (15.6) | 1.12 [-0.21, 2.40] | **.10** |
| In-community family partner/support specialist services | 6 (67%) | 19.6 (6.7) | 30.9 (16.5) | 1.17 [-0.23, 2.51] | **.10** |
| **Massachusetts Psychosis Fidelity Scale Assessment (Score Range)** |  |  |  |  |  |
| **Total Fidelity Score (0-89)** | 79.7 (4.6) | 25.0 | 3.8 (1.7) | .42 | **.06** |
| **Total Staffing and Services Score (0-34)** | 29.8 (2.2) | 20.7 (5.9) | 9.8 (2.8) | .63 | **.01 \*** |
| Supported Education and Employment (0-8) | 4.8 (2.9) | 23.3 (2.5) | 7.9 (3.2) | .51 | **.05 \*** |
| - Begins job/education search within 30 days of meeting | 5 (56%) | 23.5 (1.8) | 32.2 (16.0) | 1.27 [-0.20, 2.67] | **.09** |
| - Helps clients obtain information about Social Security, Medicaid, and other government entitlements | 3 (33%) | 30.0 (12.8) | 36.4 (14.5) | 1.60 [0.03, 3.55] | **.05 \*** |
| **Total Program Structure Score (0-19)** | 17.7 (1.0) | 52.6 | 10.9 (9.3) | .15 | .30 |
| **Total Program Procedure Score (0-36)** | 32.2 (2.0) | 25.0 | 5.6 (4.5) | .18 | .25 |
| Level of engagement practices (0-5) | 4.1 (0.8) | 18.6 (9.1) | 19.8 (10.5) | .34 | **.10 \*** |

Note. p ≤ .10 is **bolded**. \*: p ≤ .05. NA: Not applicable as low cell count.

Programs with a greater variety of outreach and engagement practices (e.g., family-friendly hours), or that were able to provide home/in-community family intervention visits (n=4), or family partner or family resource specialist services (n=6) all had marginally significantly higher family engagement rates (p=.10).

#### 3.2.2. Barriers and Engagement Strategies

Qualitative data indicated that high (n=4) and low (n=5) family engagement programs perceived similar barriers, including: 1) adverse social determinants of health; 2) scheduling difficulties; 3) adult clients’ preferences for limited family involvement; 4) family-related factors (e.g., stigma around mental health services, mismatch between treatment and family expectations, cultural and language barriers); and 5) inadequate staffing for family services (Appendix Table E.1.). Patient retention strategies described in Section 3.1.2. were also used for family engagement (Appendix Table E.2.).

Programs with high family engagement (n=4) used several distinctive approaches compared to low family engagement programs (n=5) (**Table 2**). Like high patient retention programs, high family engagement programs (75%) were more likely to provide case management and practical support for families, meeting their practical needs first to build trust and then providing psychoeducation, than low family engagement programs (40%).

All high family engagement programs strongly recommended family services upon enrollment. In contrast, only 40% of low engagement programs recommended family services at enrollment, whereas the remaining 60% of programs offered family intervention only when requested or after family crisis/conflicts emerged. For example, one team member reported, “It depends on whether the family might need the support…if I’m getting a hundred calls from a family member or they’re asking for it, we’ll talk about [providing] family support.” Relatedly, high family engagement teams were more likely to frame family services as educational rather than therapy (75% vs. 40%). One team described: “The key is approaching all families from the perspective of providing support and education rather than fixing, blaming, or pathologizing the family.”

## 4. Discussion

This mixed methods study identified engagement and retention-related programmatic factors and strategies in nine Massachusetts’ CSC programs with mean patient retention of 86% and mean family engagement of 40%. While the family engagement rate was comparable to that reported in CSC programs in research (Glynn et al., 2018) and real-world settings (Demarais et al., 2024; Oluwoye et al., 2023), the patient retention rate, which was measured over the whole program duration and included patients recently enrolled in the program, was expectedly higher than reported rates of early disengagement (Robson & Greenwood, 2022).

CSC teams with higher fidelity to evidence-based CSC components both engaged significantly more families and retained more patients than lower fidelity teams, suggesting that service quality was critical for achieving better outcomes (Bond et al., 2008; McHugo et al., 1999). This finding suggests that improved patient retention and family engagement may partly contribute to associations between CSC fidelity and better clinical outcomes (Rosenblatt et al., 2025). Higher fidelity implementation creates better conditions for sustained therapeutic relationships and meaningful service delivery to promote continued participation in treatment, maximizing treatment gains from evidence-based CSC components. Routine fidelity monitoring including engagement metrics can enhance quality improvement efforts and serve as an early patient outcome predictor (Addington et al., 2024; Breitenstein et al., 2010).

More specifically, patient retention was positively associated with fidelity to individual therapy and family services. Family interventions like psychoeducation help families reinforce their relative’s treatment involvement (Demarais et al., 2024; Oluwoye et al., 2023) and consistent family contact maintains patient’s connection with the treatment team when their motivation wanes. Additionally, family engagement was positively associated with overall fidelity to CSC services and fidelity to supported education and employment. Evidence-based supported education and employment services increase patients’ work and school participation (Rosenheck et al., 2017), potentially alleviating family stress and enabling greater family treatment participation. The association between benefits counselling and family engagement also indicates that families value guidance about considerations around their relative applying for social benefits. Taken together, these findings indicate that high-fidelity service delivery can positively impact patient retention and family engagement. While we did not collect data on program resources, achieving and sustaining high-fidelity implementation likely requires strong leadership, adequate staffing, and ongoing training and supervision (Choy-Brown et al., 2021; Durlak & DuPre, 2008; Hailemariam et al., 2019).

Programs with higher patient retention/family engagement rates reported similar treatment barriers to those with lower rates, including social determinants, patient illness-related factors, family stigma, and staffing/service limitations, all of which have been reported in prior research (Eassom et al., 2014; Selick et al., 2017). However, high and low patient retention/family engagement programs differed markedly in the engagement strategies they used. Successful programs more effectively developed approaches to mitigate barriers or enhance engagement, suggesting that engagement might depend less on the absence of specific challenges and more on the program’s ability to implement effective strategies.

Providing case management-related services was a successful strategy endorsed by both high patient retention and high family engagement programs. Case management improves treatment adherence and functioning in serious mental illness (Mueser et al., 1998; Ziguras & Stuart, 2000), especially by addressing health-related social needs (Jester et al., 2023). We found that meeting families’ practical needs functioned as a “gateway” to engagement, building trust and increasing receptivity to family psychoeducation and skills training while reducing family stress and resource burden. Case management may also facilitate family engagement by demonstrating the tangible benefits of CSC involvement, thereby reinforcing continued family contact with the team. Integrating robust case management supports or family peer specialists who can provide care navigation appears to be a high-yield engagement strategy, one that is made challenging by the lack of third-party payor reimbursement for case management activities.

Distinct predictors of either patient retention or family engagement suggest that each outcome may be optimized through different but complementary approaches. For patient retention, quantitative and qualitative findings emphasized psychotherapeutic factors that strengthen therapeutic alliance and goal alignment, including: comprehensive intake assessments with structured feedback, provider visits during hospitalization to maintain therapeutic continuity during a period vulnerable to treatment disengagement (Compton, 2005; Myers et al., 2017), and periodic treatment reviews to align services with evolving patient preferences. These factors optimize patient-centred treatment through developing a comprehensive understanding of the person needs, goals, and preferences, and maintaining the therapeutic alliance during vulnerable periods (Browne et al., 2017; Dixon et al., 2016). For family engagement, increased accessibility of the treatment team to the family, such as having capacity for proactive outreach strategies, family-friendly office hours, and flexibility to meet families at home or in the community, were more critical than other program characteristics for establishing and continuing engagement (Lucksted et al., 2018). Programs should emphasize therapeutic relationship-building and goal alignment strategies for patient retention, while prioritizing accessibility and practical supports for family engagement.

Additionally, proactively offering family services from the point of program enrollment was important, as this period represents a critical engagement window predicting sustained family treatment participation (Oluwoye et al., 2020; Oluwoye et al., 2023). Engagement of families during this period is crucial as patients and families are often confused by the onset of FEP, extremely distressed, and receptive to support from knowledgeable providers to assist them in making sense of their experiences and navigating the complexities of the treatment system (Mueser & Foo, 2024). Successful programs positioned family intervention as a standard clinical recommendation rather than an optional service, framing it as educational and supportive rather than as therapy, thereby normalizing family treatment involvement and reducing stigma. Programs may also consider using a shared decision making approach with patients during enrollment to help them determine the degree of family involvement that best fits their needs and preferences, as such methods significantly increase family participation while balancing respect for client autonomy (Davies et al., 2025; Dixon et al., 2014).

Altogether, these findings have important implications for CSC program development and quality improvement. Program services and processes can be designed with these factors and engagement strategies in mind to maximize patient retention and family engagement. Future research should examine client- and family-level predictors (Drapalski et al., 2018; Oluwoye et al., 2018, 2020; Robson & Greenwood, 2022) with program-level factors in multilevel analyses, as well as prospectively implement assertive engagement strategies for those at higher risk for disengagement.

### 4.1. Limitations

Although other studies found that patients whose families attended more sessions show greater treatment engagement themselves (Demarais et al., 2024; Oluwoye et al., 2023), patient retention and family engagement rates were not correlated across programs in this study, likely because the study lacked a continuous measure of engagement intensity (e.g., number of attended appointments). The contribution of funding and resources to fidelity is unknown, making it difficult to disentangle whether fidelity drives improved retention/engagement outcomes or whether underlying resource availability accounts for both. Relatedly, fidelity domain scores that were associated with engagement outcomes may indicate services that were more challenging to implement, thus differentiating between programs with varying capacity rather than reflecting differences in implementation quality. Given this study’s single-state, cross-sectional design, findings require replication in other FEP CSC programs, and future work should employ experimental designs to clarify causal mechanisms through which high-fidelity CSC implementation improves clinical outcomes. Finally, we lacked patient and family perspectives on engagement barriers and facilitators, which would be a valuable complement to this study’s provider perspectives (Stokes et al., 2024).

## 5. Conclusion

Higher patient retention and family engagement in FEP CSC was associated with higher-fidelity to evidence-based CSC services, suggesting treatment engagement as a potential mechanism linking high-fidelity implementation to better clinical outcomes. Programs that were more successful in retaining patients and engaging families distinguished themselves with effective engagement strategies, such as the use of case management-related supports to address common treatment barriers. Patient retention depended on psychotherapeutic factors that could strengthen therapeutic alliance and goal alignment, while family engagement relied on accessibility and proactive family service recommendation. These findings provide a data-informed approach to promoting patient and family engagement in CSC, emphasizing the need for adequate resources and training to sustain high-fidelity evidence-based services, routine fidelity monitoring incorporating engagement metrics, and tailored engagement strategies.

## Data Availability

All data produced in the present study are available upon reasonable request to the authors and IRB approval for data sharing.

## Acknowledgements

The authors thank Lisa Dixon, MD, for consultation and input during the grant proposal for this study, and all clinic staff who participated in the study.

## Author Contributions

Cheryl Y. S. Foo: *Conceptualization, Data curation, Formal analysis, Funding acquisition, Investigation, Methodology, Project administration, Supervision, Visualization, Writing – original draft;* Catherine J. Leonard: *Data curation, Formal analysis, Investigation, Project administration;* Merranda McLaughlin: *Formal Analysis, Writing – review & editing*; Kelsey A. Johnson: *Investigation, Resources, Writing – review & editing;* Dost Öngür: *Conceptualization, Funding acquisition, Supervision, Writing – review & editing;* Kim T. Mueser: *Conceptualization, Methodology, Supervision, Writing – review & editing*; Corinne Cather: *Conceptualization, Methodology, Resources, Supervision, Writing – review & editing*.

## Data Sharing Statement

Deidentified data that support the findings of this study are available on request from the corresponding author. The data are not publicly available due to privacy or ethical restrictions.

