## Supplemental Material for "A Mixed Methods Study of Program-Level Factors Influencing Patient and Family Engagement in First Episode Psychosis Coordinated Specialty Care"

Table of Contents for Appendices

### **Appendix A. Massachusetts Psychosis Fidelity Scale (MAPS) Categories, Domains, and Domain Criteria Assessed in this Study**

| **Category (Score Range)** | **Domain** | **Number of Criteria/Total Domain Score** |
| --- | --- | --- |
| **Staffing and Services (0-34)** | Team Leader Role | 3 |
|  | Individual Therapy | 2 ^a^ |
|  | Family Services | 7 ^a^ |
|  | Medication Management | 6 |
|  | Health Management | 5 |
|  | Supported Education and Employment | 8 |
|  | Case Manager/Care Coordinator | 3 |
| **Program Structure (0-19)** | Patient-to-Provider Ratio | 2: <20:1; 1: 20:1 to 40:1; 0: >40:1 |
|  | Team Meeting Frequency | 2: Weekly; 1: 1-3 times per month; 0: <1x per month |
|  | Eligibility Criteria | 2: ≥ 80%; 1: 60-70%; 0: <60% meeting program eligibility for first-episode psychosis |
|  | Program Length | 2: ≥2 years; 1: 1-2 years; 0: Under 1 year |
|  | Crisis and Safety | 4 |
|  | Care Coordination with Inpatient Services | 7 |
| **Program Procedures (0-36)** | Outreach and Referrals | 3 |
|  | % clients seen within 2 weeks of referral | 2: >60%; 1: 20-60%; 0: <20% |
|  | Level of engagement practices | 5 |
|  | Clinical & Psychosocial Needs Assessment | 11 |
|  | Individualized treatment plan | 6 |
|  | Quality Improvement | 2 |
|  | Cultural Responsiveness | 3 |
|  | Psychoeducation | 4 |

^a^ For this study, engagement benchmark criteria under the “individual therapy” and “family services” domains were removed to avoid confounding with primary outcomes.

### **Appendix B. Group Interview Guide**

A. **Briefing and Consenting Process [2 mins]**
B. **Patient Retention and Engagement [30 mins]**

*Let’s start with patient retention first. It’s an important issue because we want our clients to get the most benefit out of early intervention for psychosis, which is recommended to be at least 2-3 years but anywhere from 15-50% dropout by the first year of enrolment when they can benefit from continuing to be in treatment.*

*Following SAMHSA definitions, a client is considered “disengaged” if they are “lost due to follow-up or otherwise discontinued services not due to reasons for planned discharge (e.g., recovery, or completing/graduating from time-limited treatment), hospitalization, death, or transfer to criminal legal system.” So, we are specifically focusing on the challenges of retaining individuals who would benefit from continuing to be treatment.*

***Operationalization of Patient ‘Retention’ and ‘Engagement’:***

1. **Broadly, what are your program’s expectations of “staying in treatment” for a client?**
   a. E.g. minimum time in treatment, active participation in number of services (minimum requirements), legitimate reasons for disengagement, maximum no. of months not receiving treatment/missed sessions, reengagement after gap in treatment
   b. What are the criteria for discharging someone from your program?
   c. In your program, is there a minimum number or types of coordinated specialty care components that a client should be “engaged in” in order to continue to participate in the program (vs. patient preference)?
      i. Can a patient who refuses to take medications or see the psychiatrist still be retained in the program?
   d. What is the minimum duration of time someone can not receive services and still be in the program?
   e. What will count as ‘early/premature discharge’ (e.g., disengage by 6/9/12mo)?

*Program Metrics and Self-Rated Satisfaction:* **[Use data from MAPNET/program leader survey: “Your program reported X% of clients retained in services for at least one year.”]**

2. **How good do you think your program’s retention rates are? (Rate 1-5).**

***Barriers, Facilitators, and Strategies for Patient Engagement:***

3. **What are challenges your program has faced in keeping clients involved in the services your program provides?**
   a. *[Prompt for individual, organizational, systemic barriers]*
   b. *What are some of the common reasons for client dropping out of the program?*
   c. Are there components of coordinated specialty care that clients have more difficulty with that contribute to dropout? [Client factor]
   d. Are there particular organizational or system barriers that make it hard to retain certain clients (e.g., billing requirements)?
4. **What strategies, approaches, or workflow have you found worked well for retaining clients in your treatment program?**
   a. Gather information about program’s philosophy and approach towards patient engagement: e.g. how are they oriented to CSC program and FEP treatment; core principles of FEP treatment e.g., respective patient preference while maintaining therapeutic alliance
   b. Is there a program orientation? What is included in the program orientation? [Share any materials that they use with clients and families] When does the orientation take place? (e.g., is it after a diagnostic evaluation and/or team discussion?) When do they get started with individual and family services after the program orientation? Who is responsible for contacting the client/family and conducting the program orientation?
   c. **What specific strategies have you found worked well for keeping clients involved in:**
      i. Accepting and adhering to medications?
      ii. Individual psychotherapy?
      iii. Attending joint family psychoeducation and support sessions?
      iv. SEE?
      v. Other CSC treatment components?
C. **Family Engagement [25 mins]**

*The other piece that we are interested in is family engagement in your program. Again, we know that family involvement and support can help improve patient’s adherence to treatment, but rates of family participation in first-episode treatment services similarly range only from 23-48% in the United States.*

***Program’s philosophy and approach to family involvement and family services:***

5. **Some programs believe in always including the family in the client’s treatment and recommending family-based intervention (e.g. psychoeducation program) for all clients, while others think families should choose if they want to be involved or not (e.g., “opt-in” approach for certain family interventions). Which way does your program lean?**
   a. Why has it been practiced this way?
   b. Gather key information about program’s philosophy and approach to family involvement and family-based services: e.g., when and for what purposes are families typically engaged and involved; different policies for family involvement based on age/type of client; at what point of treatment are families involved/contacted; when are family-based services introduced to clients/families, and how are these families identified
   c. When families are the referral source and first point of contact, how does the team work with the family to engage the client? Can families participate in treatment if the client is not engaged in treatment?
   d. Can families participate in treatment if the client does not attend family sessions, but are involved in other aspects of the program? Typically, what % of family sessions do the clients jointly attend?

*Program Metrics and Self-Rated Satisfaction:* **[Use data from MAPNET/program leader survey: “Your program offers these family services [list], and reported X% of clients who live with/are in contact with family are involved in family services.”]**

6. How good do you think your program’s family engagement rates are?

***Barriers, Facilitators, and Strategies for Family Engagement***

7. **What are the challenges your program has faced in involving families in treatment?**
   a. [Prompt for individual, organizational, systemic barriers]
   b. What are some of the reasons for patient/family refusal?
8. **What ways have you seen made it easier for families to stay involved in your program?**
   c. E.g. outreach and communication, family day, monthly/annual family meetings about patient care/treatment plan?
D. **Recommendations for Patient and Family engagement**
9. **What are 1-2 best suggestions you have to keep patients and families involved in treatment?**
10. **What extra supports or training does your program need to make those improvements?**
E. **Summary and thank you [2 mins].**

### **Appendix Table C.1.** Characteristics of First-Episode Psychosis Coordinated Specialty Care Programs in Massachusetts (N=9)**.**

| ***Program Characteristic*** | **N (%) / M (SD)** | **A** | **B** | **C** | **D** | **E** | **F** | **G** | **H** | **I** |
| --- | --- | --- | --- | --- | --- | --- | --- | --- | --- | --- |
| Total enrolled clients served in 2022-2023 | 65 (33) | 45 | 25 | 33 | 201 | 61 | 31 | 147 | 19 | 19 |
| Time since program established (years) | 6.7 (SD) | 8 | 3 | 7 | 4 | 20 | 4 | 11 | 2 | 1 |
| Implements NAVIGATE CSC Model | 7 (78) |  | X | X | X | X | X |  | X | X |
| Age served | - | 16-30 | 13-35 | 17-40 | <30 | 14-35 | 15-40 | 18-30 | 13-21 | 16-40 |
| High patient to FTE Staff ratio (>20:1) | 10.9 (6) |  |  |  | X | X |  |  |  |  |
| Able to conduct home/community visits | 4 (44) | X |  | X |  |  |  |  | X | X |
| ***Family Services Characteristics*** |  |  |  |  |  |  |  |  |  |  |
| Designated family services coordinator/lead | 6 (67) | X^a^ | X |  | X | X |  |  | X^a^ | X^a^ |
| High patient to FTE Family Clinician ratio (>40:1) | 75.8 (80) |  |  | X | X | X |  | X |  |  |
| Ability to sufficiently bill for family services | 5 (56) |  | X | X | X |  | X |  |  | X |
| Able to hold family sessions outside of regular office hours | 7 (78) | X |  |  | X | X | X | X | X | X |
| Provides NAVIGATE Family Education and Support Program | 7 (77) | X | X | X | X | X | X |  |  | X |
| Provides McFarlane Multifamily Group for Psychosis | 3 (33) |  |  | X |  | X |  | X |  |  |
| Provides family support group | 4 (44) | X |  |  | X |  | X | X |  |  |
| Provides family partner/family peer services | 3 (33) | X |  |  |  |  | X |  |  | X |
| Conducts treatment reviews with family members | 7 (8) | X | X |  |  | X | X | X | X | X |
| Patients’ family members receive at least 6 months of family intervention | 4 (44) | X | X |  |  |  |  | X |  | X |
| Family intervention is conducted at least fortnightly in the first 6 months | 5 (56) | X | X |  |  | X | X | X |  |  |
| ≥50% of family sessions conducted with client present | 4 (44) |  | X |  |  | X |  | X |  | X |

^a^ Team leader plays role of family services coordinator/lead, which is also consistent with NAVIGATE model’s recommendations.

### **Appendix Table D.1.** Program-Level Factors Affecting Patient Retention Rates (full list of predictors explored).

| **Program-Level Factor** | **N (%) / M (SD)** | **Observed mean patient retention rate at min *x* (SD)** | **B (SE)** | **Effect size**  ***g* [95% CI] / R^2^** | **p-value** |
| --- | --- | --- | --- | --- | --- |
| Estimated distance travelled (1-10 miles vs. 11-20 miles) | 4 (44%) | 78.4 (15.2) | -12.9 (8.1) | -0.95 [-2.19, 0.34] | .15 |
| ***Services Provided*** | | | | | |
| In-clinic/telehealth family intervention | 8 (89%) | 57.9 | 31.2 (9.1) | 3.23 [0.69, 5.65] | **.01 **** |
| In-community family intervention visits | 4 (44%) | 79.5 (15.2) | 13.8 (7.8) | 1.05 [0.27, 2.31] | **.**12 |
| Peer support services | 8 (89%) | 57.9 | 31.2 (9.1) | 3.23 [0.69, 5.65] | **.01 **** |
| Recreational and support groups | 6 (67%) | 80.7 (8.4) | 7.3 (9.5) | 0.48 [-0.79, 1.72] | .47 |
| Case management | 7 (78%) | 83.1 (10.4) | 30.0 (19.9) | 1.07 [-0.46, 2.54] | .18 |
| Housing support services | 6 (67%) | 84.6 (15.2) | -2.9 (9.9) | -0.19 [-1.41, 1.06] | .78 |
| ***Massachusetts Psychosis Fidelity Scale Assessment (Score Range)*** | | | | | |
| **Total Fidelity Score (0-89)** | 79.7 (4.6) | 97.3 | 0.8 (1.0) | .08 | .44 |
| **Total Staffing and Services Score (0-34)** | 29.8 (2.2) | 84.9 (10.7) | 1.1 (2.3) | .03 | .65 |
| Team Leader (0-3) | 3 (0.0) | NA | NA | NA | NA |
| Individual Therapy (0-2) | 2.8 (0.4) | 57.9 | 31.2 (9.9) | .62 | **.01 **** |
| - Provides evidence-based individual therapy | 9 (100%) | NA | NA | NA | NA |
| - Trained in evidence-based individual therapy | 8 (89%) | 57.9 | 31.2 (9.9) | 3.23 [0.69, 5.65] | **.01 **** |
| Prescriber (0-6) | 6 (0.0) | NA | NA | NA | NA |
| Health Management (0-7) | 6.7 (1.0) | 93.5 | -3.0 (4.8) | .05 | .56 |
| Family Services (0-5) | 4.2 (1.0) | 69.9 (10.4) | 11.5 (2.7) | .72 | **<.01 **** |
| - Provides evidence-based family interventions | 8 (89%) | 57.9 | 31.2 (9.1) | 3.23 [0.69, 5.65] | **.01 **** |
| - Trained in evidence-based family interventions | 8 (89%) | 57.9 | 31.2 (9.1) | 3.23 [0.69, 5.65] | **.01 **** |
| - >60% families involved in initial assessment | 6 (67%) | 81.7 (10.1) | 5.9 (9.7) | 0.38 [-0.87, 1.61] | .56 |
| - >60% families have a signed ROI for family-team contact | 9 (100%) | NA | NA | NA | NA |
| - >60% contacted quarterly by one team member | 7 (78%) | 75.9 (0.17) | 12.5 (10.2) | 0.87 [-0.62, 2.31] | .26 |
| Supported Education and Employment (0-8) | 4.8 (2.9) | 93.9 (4.8) | 0.1 (1.7) | .00 | .95 |
| Case Management (0-3) | 2.7 (0.5) | 88.7 (11.3) | -4.6 (9.8) | .03 | .65 |
| **Total Program Structure Score (0-19)** | 17.7 (1.0) | 57.9 | 7.5 (4.1) | .33 | **.10 *** |
| Adequate patient-to-provider ratio (<20:1 vs. 20:1 to 40:1) | 7 (78%) | 83.3 (14.1) | 10.2 (10.6) | 0.69 [0.77, 2.11] | .36 |
| Team meeting frequency (1-3 times/month vs. weekly) | 3 (33%) | 77.0 (19.7) | 12.9 (8.6) | 0.94 [-0.41, 2.24] | .18 |
| 80% of patients meeting eligibility criteria | 8 (89%) | 57.9 | 31.2 (9.9) | 3.23 [0.69, 5.65] | **.01 **** |
| ≥2 years program duration | 9 (100%) | NA | NA | NA | NA |
| Crisis Management (0-4) | 3.9 (0.3) | 93.5 | -8.9 (14.5) | .05 | .56 |
| Coordination with Inpatient Services (0-7) | 6.6 (0.5) | 80.7 (8.4) | 13.3 (4.0) | .65 | **.02 **** |
| - Inpatient unit contacted | 8 (89%) | NA | NA | NA | NA |
| - Visit with patient on inpatient unit | 5 (56%) | 80.7 (8.4) | 13.3 (4.0) | 2.11 [0.37, 3.78] | **.02 **** |
| - Communicate with family about admission | 8 (89%) | NA | NA | NA | NA |
| - Are involved in discharge planning process | 8 (89%) | NA | NA | NA | NA |
| - Receive/obtain a hospital discharge summary | 8 (89%) | NA | NA | NA | NA |
| - Schedule outpatient appointment prior to discharge | 8 (89%) | NA | NA | NA | NA |
| - Face-to-face contact with FEP provider within 2-weeks post-discharge | 8 (89%) | NA | NA | NA | NA |
| **Total Program Procedure Score (0-36)** | 32.2 (2.0) | 97.2 | 1.3 (2.4) | .04 | .61 |
| Outreach and referrals (0-3) | 2.8 (0.4) | 77.6 (27.8) | 10.3 (10.6) | .12 | .36 |
| >60% clients have appointment within two weeks from referral | 5 (56%) | 90.0 (10.0) | -7.9 (8.9) | -0.54 [-1.71, 0.68] | .40 |
| Level of engagement practices (0-5) | 4.1 (0.8) | 86.5 (15.2) | -4.26 (6.1) | .06 | .51 |
| Clinical and psychosocial needs assessment (0-11) | 10.4 (0.7) | 57.9 | 15.9 (3.3) | .77 | **<.01 **** |
| - Individual and family psychiatric history at intake | 8 (89%) | 57.9 | 31.2 (9.1) | 3.23 [0.69, 5.65] | **.01 **** |
| - Feedback is offered to client (and family) | 5 (56%) | 75.8 (14.6) | 17.6 (6.6) | 1.58 [0.12, 2.97] | **.03 **** |
| Individualized treatment plan (0-6) | 5.6 (0.7) | 57.9 | 8.5 (6.0) | .22 | .20 |
| Quality improvement processes (0-2) | 1.2 (0.4) | 90.4 (7.3) | -23.4 (6.6) | .65 | **<.01 **** |
| - Reviewing feedback from clients and families | 8 (89%) | 90.4 | -5.4 (14.7) | -0.35 [-2.19, 1.52] | .72 |
| - Setting team-based QI goals at least once per year | 3 (33%) | 91.0 (8.0) | -16.2 (7.8) | -1.31 [-2.68, 0.12] | **.08 *** |
| Culturally responsive services (0-3) | 2.8 (0.4) | 95.4 (2.6) | -12.6 (10.2) | .06 | .26 |
| Psychoeducation (0-4) | 3.8 (0.4) | 75.9 (0.2) | -12.6 (10.2) | .18 | .26 |

*Note.* * p ≤ .10, ** p ≤ .05. NA: Not applicable as low cell count.

### **Appendix Table D.2.** Program-Level Factors Affecting Family Engagement Rates (full list of predictors explored).

| **Factor** | **N (%) / M (SD)** | **Observed mean family engagement rate at min *x* (SD)** | **B (SE)** | **Effect size**  ***g* [95% CI] / AR^2^** | **p-value** |
| --- | --- | --- | --- | --- | --- |
| Adequate patient-to-family provider ratio (<40:1) | 5 (56%) | 50.7 (30.5) | -23.7 (17.0) | -0.83 [-2.05, 0.44] | .21 |
| High leadership prioritization of family services (5) | 5 (56%) | 21.1 (6.3) | 34.3 (14.1) | 1.45 [0.03, 2.79] | **.05 **** |
| Designated family service coordinator | 6 (67%) | 29.0 (19.3) | 16.6 (19.2) | 0.54 [-0.74, 1.79] | .41 |
| ≥50% of family intervention sessions conducted with client present | 4 (44%) | 49.5 (35.6) | 17.0 (18.1) | 0.56 [-0.66, 1.74] | .38 |
| Ability to schedule family sessions outside office hours | 7 (78%) | 46.4 (27.1) | 28.4 (20.3) | 1.00 [-0.52, 2.45] | .20 |
| Adequate billing for family services | 5 (56%) | 38.4 (14.6) | 3.2 (19.1) | 0.10 [-1.07, 1.26] | .87 |
| Families receive ≥6 months of family intervention | 5 (56%) | 45.7 (32.6) | 12.5 (18.6) | 0.40 [-0.80, 1.57] | .52 |
| Weekly family sessions in first 6 months of program | 5 (56%) | 35.0 (13.4) | -11.6 (18.7) | -0.37 [-1.54, 0.83] | .55 |
| Estimated distance travelled (1-10 miles vs. 11-20 miles) | 4 (44%) | 49.5 (36.5) | -16.9 (18.1) | -.56 [-1.74, 0.66] | .38 |
| ***Services Provided*** |  |  |  |  |  |
| In-clinic/telehealth family intervention | 8 (89%) | 52.6 | -14.1 (29.9) | -0.44 [-2.3, 1.43] | .65 |
| In-community family intervention visits | 4 (44%) | 27.1 (15.2) | 29.4 (15.6) | 1.12 [-0.21, 2.40] | **.10 *** |
| In clinic/telehealth family peer support services | 5 (56%) | 27.0 (15.8) | 23.7 (17.0) | 0.83 [-0.44, 2.05] | .21 |
| In-community family partner/support specialist services | 6 (67%) | 19.6 (6.7) | 30.9 (16.5) | 1.17 [-0.23, 2.51] | **.10 *** |
| Case management | 7 (78%) | 16.8 (6.7) | -30.0 (19.9) | -1.07 [-2.54, 0.46] | .18 |
| Housing support services | 6 (67%) | 29.6 (19.1) | 18.5 (17.9) | 0.51 [-0.76, 1.76] | .44 |
| ***Massachusetts Psychosis Fidelity Scale Assessment (Score Range)*** | | | | | |
| **Total Fidelity Score (0-89)** | 79.7 (4.6) | 25.0 | 3.8 (1.7) | 0.42 | **.06 *** |
| **Total Staffing and Services Score (0-34)** | 29.8 (2.2) | 20.7 (5.9) | 9.8 (2.8) | 0.63 | **.01 **** |
| Team Leader (0-3) | 3 (0.0) | NA | NA | NA | NA |
| Individual Therapy (0-2) | 2.8 (0.4) | 52.6 | -14.1 (29.9) | .03 | .65 |
| Prescriber (0-6) | 6 (0.0) | NA | NA | NA | NA |
| Health Management (0-7) | 6.7 (1.0) | 50.0 | -3.7 (10.0) | .02 | .72 |
| Family Services (0-5) | 4.2 (1.0) | 29.6 (20.8) | 10.1 (9.7) | .14 | .33 |
| Supported Education and Employment (0-8) | 4.8 (2.9) | 23.3 (2.5) | 7.9 (3.2) | .51 | **.05 **** |
| - SEE specialist has BA-level training | 8 (89%) | NA | NA | NA | NA |
| - Provides varied supports at work, school, and in the community | 7 (78%) | 25.0 | 21.3 (29.0) | 0.68 [-1.21, 2.51] | .49 |
| - Contacts >80% of clients within 30 days of enrolment | 5 (56%) | 31.9 (15.1) | 18.6 (19.3) | 0.62 [-0.70, 1.89] | .37 |
| - Meets with clients monthly for as long as client wants/needs | 6 (67%) | 23.3 (2.5) | 27.1 (20.4) | 0.94 [-0.57, 2.40] | .23 |
| - Begins job/education search within 30 days of meeting | 5 (56%) | 23.5 (1.8) | 32.2 (16.0) | 1.27 [-0.20, 2.67] | **.09 *** |
| - Seeks competitive employment or educational placements | 6 (67%) | 22.8 (1.7) | 27.8 (20.2) | 0.98 [-0.55, 2.43] | .22 |
| - Visits schools/employers | 3 (33%) | 33.9 (14.3) | 25.8 (17.9) | 0.92 [-0.46, 2.23] | .20 |
| - Helps clients obtain information about Social Security, Medicaid, and other government entitlements | 3 (33%) | 30.0 (12.8) | 36.4 (14.5) | 1.60 [0.03, 3.55] | **.05 **** |
| Case Management (0-3) | 2.7 (0.5) | 37.1 (21.6) | 4.5 (20.1) | .01 | .83 |
| **Total Program Structure Score (0-19)** | 17.7 (1.0) | 52.6 | 10.9 (9.3) | .15 | .30 |
| Adequate patient-to-provider ratio (<20:1 vs. 20:1 to 40:1) | 3.8 (0.4) | 31.9 (15.1) | -12.3 (19.7) | -0.39 [-1.63, 0.87] | .55 |
| Team meeting frequency (1-3 times/month vs. weekly) | 3 (33%) | 29.9 (20.7) | 17.2 (18.0) | 0.57 [-0.66, 1.75] | .37 |
| 80% of patients meeting eligibility criteria | 8 (89%) | 52.6 | -14.1 (29.9) | -0.44 [-2.29, 1.43] | .65 |
| ≥2 years program duration | 9 (100%) | NA | NA | NA | NA |
| Crisis Management (0-4) | 3.9 (0.3) | 50.0 | -11.1 (30.0) | .02 | .72 |
| Coordination with Inpatient Services (0-7) | 6.6 (0.5) | 19.2 (6.3) | 31.0 (18.2) | .32 | .14 |
| **Total Program Procedure Score (0-36)** | 32.2 (2.0) | 25.0 | 5.6 (4.5) | .18 | .25 |
| Outreach and referrals (0-3) | 2.8 (0.4) | 38.8 (19.5) | 1.7 (22.9) | .00 | .94 |
| >60% clients have appointment after two weeks from referral (2) | 5 (56%) | 27.0 (15.8) | 23.7 (17.0) | 0.83 [-0.44, 2.10] | .21 |
| Level of engagement practices (0-5) | 4.1 (0.8) | 18.6 (9.1) | 19.8 (10.5) | .34 | **.10 *** |
| Clinical and psychosocial needs assessment (0-11) | 10.4 (0.7) | 52.6 | 2.19 (13.9) | .00 | .88 |
| Individualized treatment plan (0-6) | 5.6 (0.7) | 52.6 | -4.68 (13.8) | .02 | .74 |
| Quality improvement processes (0-2) | 1.2 (0.4) | 40.6 (29.7) | -2.3 (22.9) | .00 | .92 |
| Culturally responsive services (0-3) | 2.8 (0.4) | 37.5 (17.7) | 3.4 (22.9) | .00 | .89 |
| Psychoeducation (0-4) | 3.8 (0.4) | 18.0 (8.4) | 28.4 (20.3) | .22 | .20 |

*Note.* * p ≤ .10, ** p ≤ .05. NA: Not applicable as low cell count.

### **Appendix Table E.1.** Perceived Engagement Barriers across Higher and Lower Patient Retention and Family Engagement Programs.

| **Code** | **Number of teams endorsed (N=9)** | **Higher patient retention team**  **(n=6)** | **Lower patient retention team (n=3)** | **Higher family engage-ment team (n=4)** | **Lower family engage-ment team (n=5)** | **Quotes** |
| --- | --- | --- | --- | --- | --- | --- |
| ***Barriers to both Patient Retention and Family Engagement*** | | | | | | |
| Adverse social determinants of health (e.g., housing instability, transportation difficulties) | 7 | 83% | 67% | 75% | 80% | *“One of the barriers that patients have faced that have led to disengagement, pre-COVID was transportation. But with [telehealth], some of my patients will disengage because they don’t have access to a computer.” (Team D)* |
| ***Specific Barriers to Patient Retention*** | | | | | | |
| Patient illness-related factors (e.g., lack of insight, substance use, poor medication adherence) | 8 | 83% | 100% | - | - | *“In our experience, someone in early adulthood oftentimes do not want to accept or connect with the reality of what's changed in their lives and accept [schizophrenia] as a major mental illness.” (Team B)* |
| Limited staffing and staff turnover | 2 | 17% | 33% | - | - | *“We don't have the funding for full-time staff for any of our positions, aside from the peer specialist and the employment and education specialist, which we're working on hiring… We're using our fee-for-service clinicians in the clinic to get trained who want to do this work. But that means that there's a limited number of hours a week that they're able to provide for.” (Team B)* |
| ***Specific Barriers to Family Engagement*** | | | | | | |
| Difficulty scheduling families during typical working hours | 6 | - | - | 75% | 60% | *“Just the practical challenges of parents or caregivers who are working 9:00 to 5:00, and those are the times that we're available” (Team A)*  *“The top barrier is scheduling around family’s work schedules. They have a 9 to 5 or even 7 to 7.” (Team F)* |
| Adult client’s preference for limited family involvement | 5 | - | - | 50% | 60% | *“The 18-year-olds want some independence. They'll kind of limit us to how much they want some family members to engage.” (Team H)*  *“Most of the [young adults] they’re like, “I don’t want my family involved in this at all,” so getting families to participate has been historically challenging.” (Team B)* |
| Family-related factors |  |  |  |  |  |  |
| Family limited understanding about and stigma around mental illness/ psychosis | 4 | - | - | 50% | 40% | *“I think certainly stigma - there can be some skepticism around mental health treatment, unfortunately, still at this point.” (Team G)*  *“The family was against most medical recommendation. Even when we provided psychoed around psychosis, the family just was in denial.” (Team D)* |
| Family expectations of recovery and their role in treatment | 3 | - | - | 50% | 20% | *“[When they enter treatment, some families believe] it stops being a family problem. It starts being an individual problem. Then it's back to, "It's a moral character issue. They’re lazy. They're difficult to motivate." And it gets hard for the team to rally the family’s support” (Team I)* |
| Complex families (e.g., families with mental health issues, high conflict families) | 3 | - | - | 25% | 40% | *“There are some instances where it's difficult for the family to be involved - live having family members who have dealt with psychosis themselves or in their parents, or there might be some difficult family dynamics and history.” (Team G)* |
| Cultural and language barriers | 1 | - | - | 25% | 0% | *“Language can complicate things a bit… different languages spoken within the family themselves.” (Team I)* |
| Limited staffing for family services | 3 | - | - | 25% | 40% | *“We do not have a family clinician specifically, and I do the family work as an individual therapist. The goal is to eventually hire a family clinician so that we are able to follow through with fidelity. (Team I)* |

### **Appendix Table E.2.** Commonly Endorsed Engagement Strategies for Patient Retention and Family Engagement.

| **Code** | **Number of teams endorsed (N=9)** | **Higher patient retention team**  **(n=6)** | **Lower patient retention team (n=3)** | **Higher family engage-ment team (n=4)** | **Lower family engage-ment team (n=5)** | **Quotes** |
| --- | --- | --- | --- | --- | --- | --- |
| Importance of building trust and rapport | 9 | 100% | 100% | 100% | 100% | *“What works for me is just meeting clients where they are and listen and not rushing through the process of recovery. You just want to be respectful and build trust.” (Team I)* |
| Practicing person-centered choice and preference | 7 | 67% | 100% | 75% | 80% | *“We say any treatment decision is up to the individual. We're not going to force you to do anything… We try to just say that off the bat like, ‘Here's all this information. We understand it's an overload, but we can take it baby steps. We don't need to jump right into all of this. We're going to take it at your pace.’”*  *(Team F)* |
| Adopting whole-team, coordinated approach to engaging patients (e.g., treatment team coordinates on what services have been underutilized or should be recommended, introduce new CSC services through provider who has most engagement with patient, or by utilizing each other’s sessions/milieu activities) | 8 | 100% | 67% | 75% | 100% | *“We've had a few patients I can think of who maybe were really engaged with their individual therapist, but maybe less with psychopharm or vice versa. And so, working as a team to kind of encourage people to participate in all components of treatment.”(Team E)* |
| Use of program orientation/”meet-and-greet” to set treatment expectations   - Most teams typically prioritize clinical components (medication and therapy), and minimum engagement in 1-2 CSC components | 8 | 83% | 100% | 100% | 80% | *“The Individual Resiliency Training clinician will meet with them one-on-one and really in-depth talk about the program, kind of what it entails, what each provider kind of does, and then go from there… It’s a requirement to meet with a prescriber even if they don’t want to take meds.” (Team B)*  *“So our participants should attend at least two modalities. So for instance, the minimum having the individual therapist, and the prescriber. But we do encourage, and most stay engaged once they do other services that we have, whether that's our supported education and employment specialist, peer specialist support, and then finally our milieu when they're ready.” (Team A)* |
| Assertive outreach practices (i.e, high-touch, high-frequency contact) | 6 | 67% | 67% | 100% | 40% | *“We’re tenacious with outreach. When people are missing appointments or not getting back to us, we'll do phone calls-- phone calls from the clinician, phone calls from our front desk administrator, emails, gateway messages, letters-- all of it geared toward keeping people engaged in treatment.” (Team E)*  *“We have the staffing to continue to call, to continue to write another email, to follow up and ask the family about scheduling. And not put it on the family to reach out and schedule. We’re able to be that forward and try and get families engaged.”*  *(Team F)* |
| Use of multiple treatment delivery formats   - Home/in-community visits | 5 | 50% | 67% | 50% | 60% | *“I feel like the best engagement has been out in the community. I think that it's difficult to - I know for myself personally - want to come into a building that also is right next to an inpatient unit. I think the environment matters. So I think that it's important to meet them where they feel most comfortable.” (Team I)* |
